# Evaluating pregnant women’s selenium intake through dietary assessment in Accra, Ghana: a feasibility study

**DOI:** 10.1101/2023.01.18.23284731

**Authors:** Amy Amuquandoh, Marie Aluko, Ofori-Attah Ebenezer, Godfred Egbi, Kwame Adu-Bonsaffoh

**Author notes:** These authors contributed equally to this work (Amy Amuquandoh, Marie Aluko, Kwame Adu-Bonsaffoh).

## Abstract

**Objective:** Dietary selenium (Se) deficiency is associated adverse pregnancy outcomes. We sought to determine the feasibility of: (1) assessing dietary Se consumption via administration of a food frequency questionnaire (FFQ) to pregnant women attending the antenatal clinic of Korle Bu Teaching Hospital (KBTH) in Accra, Ghana and (2) quantifying total Se intake.

**Methods:** In this prospective observational cohort study, participants were recruited via purposive sampling and face-to-face data collection was undertaken using structured FFQs. English-speaking pregnant women who were ≥ 18 years old were eligible to participate. Questions of feasibility were assessed and the average total Se intake in the population was analyzed.

**Results:** One hundred study participants met the inclusion criteria out of 117 eligible. It was feasible to administer the surveys in the clinic waiting room. The average total Se intake in this population of pregnant women was 90.4 μg/day ±50.0 μg, which is higher than the National Institute of Health recommended dietary allowance (RDA) of 60μg/day for pregnant women.

**Conclusion:** FFQ assessment of Se consumption and resource evaluation were deemed feasible in the KBTH setting. Participants reported an average Se intake above the RDA. Future studies can examine the influence of Se intake on pregnancy outcomes.

## Introduction

Pregnant women in low and middle income countries (LMIC) are susceptible to multiple micronutrient deficiencies as a consequence of globalization and inequitable resource allocation. ^1^ Micronutrient deficiencies are capable of exerting a profound and negative effect on maternal health, fetal development and pregnancy outcomes. ^2,3^

Selenium (Se) is an essential micronutrient that humans receive predominantly through diet.^4^ Its deficiency is relatively common among pregnant women because of the increased demand for Se during gestation.^5^ In general, maternal nutrition prior to and during pregnancy strongly influences the risk of pregnancy complications. ^5^ The recommended dietary allowance (RDA) of Se for a pregnant woman is 60 μg/day (compared to 55 μg/day for non-pregnant women), and the tolerable upper level intake is 400 μg/day per the NIH.^6^ Over the past few decades, studies have described the antioxidant role of Se in placental development and in gestational fetal weight gain. ^7–10^ A deficiency in Se causes altered placental function and low fetal birth weight.

Studies show that mean maternal Se concentrations are lower in mothers who have adverse pregnancy outcomes, as described in various populations irrespective of geographic location.^11–13^ In Nigeria, a case-control study examining the relationship between serum Se deficiency and preeclampsia demonstrated that about 17.2% of the pregnant population in the control group was Se deficient.^14^ We would expect to observe the same cross-sectional relationship in pregnant women of Ghana, however, it is not currently known whether this is the case. A 2010 study done in Ghana demonstrated that serum Se levels for male and female, rural (Brekusu) and urban (Accra), subjects were within reference ranges, though this effect was not directly studied in the pregnant population.^15^ Furthermore, the process of collecting dietary recall data from pregnant women as a means of exploring this relationship has rarely been studied. Dietary Se concentrations, while less strongly predictive, is an appropriate proxy for serum Se levels. ^16^

The goal of this study is to determine the feasibility of collecting dietary Se information, via a food frequency questionnaire (FFQ), among pregnant women attending the Korle Bu Teaching Hospital (KBTH) antenatal clinic in Accra, Ghana. We hypothesize that if this approach is feasible, the survey data we collect should allow us to approximate the average dietary Se intake in this pregnant population. KBTH is a setting that is likely suited to carry out this study because it is a teaching hospital, has a high-volume antenatal clinic, and skilled health worker staff. Given the importance of Se in pregnancy and the limited data in Ghana, the findings of this feasibility study will not only evaluate the capacity of KBTH to administer dietary surveys to its pregnant population, but also provide local evidence concerning the burden of Se deficiency in pregnant women attending KBTH in Accra, Ghana.

## Methods

### Study Design & Site

This feasibility study, a prospective observational cohort design, explored the challenges and opportunities presented by administering dietary recall surveys to pregnant women in Accra, Ghana. Over a period of 3 days from May 11^th^, to May 13^th^, 2022, a team of research assistants (RA) administered FFQs to pregnant women at the antenatal clinic of the KBTH in Accra, Ghana. The antenatal clinic attends to about 150 pregnant women each day. Most pregnant women presenting to the clinic are referred high risk cases from peripheral hospitals in the Greater Accra Region and the neighboring regions.^17^

### Study Population & Selection

Our study population was drawn from pregnant women who attended the KBTH antenatal clinic. Fluency in English was a requirement to participate in the study as required by the University of North Carolina IRB. Purposive sampling was used to select participants. All adult pregnant women ≥ age of 18 attending the clinic were eligible for the study, regardless of health status or gestational age.

### Sample size determination

As this is a feasibility and hypothesis-generating study, a power calculation was not indicated. We selected a sample size of 100 pregnant women in order to estimate the average Se intake in this population; we are not able to calculate power since we have no current data from studies establishing an approximation of Se concentration in pregnant women in Ghana.

### Study Procedures

Participants arrived at the clinic and waited to be seen on a first-come-first-serve basis between the hours of 8:00 a.m-2:00 p.m. We administered a paper version of the REDCap Global (NC TraCS Institute) FFQ adapted from the one used for the Environment, Perinatal Outcomes, and Children’s Health (EPOCH) study done at UNC (Manuck, PI). We sought to answer questions such as: “How long does it take to administer a survey? Which faculty are best suited to administer the survey? How many questionnaires can we administer in one day?” (Supplemental information) We administered the FFQ captured self-reports of dietary Se intake by asking women to report which foods, from a list of about 60 food items or prepared dishes they had eaten during the past month. We then estimated the concentrations of maternal Se intake in μg/day and compared them to the 60 μg/day RDA for pregnant women.^6^ To adapt the FFQ for use in this locale, Ghanaian foods were substituted as appropriate (Table 1).

**Table 1.** Se concentration of commonly consumed Ghanaian foods determined by ICP-MS

| Name of Sample | Se Concentration (mg/kg) |
| --- | --- |
| Kenkey | 0.03 |
| Fufu | 0.06 |
| Yam | 0.01 |
| Goat meat | 1.10 |
| Pork | 0.90 |
| Brown rice | 0.05 |
| Bambara beans | 0.07 |
| Red beans | <0.01 |
| Kontomire leaves | 0.08 |
| Hausa koko (millet) | 0.12 |
| Onions | 0.02 |
| Garlic | 0.02 |
| Keta School Boys Fish (anchovies) | 0.91 |
| Jollof rice | 0.05 |
| Waakye (rice and beans) | 0.04 |
| Banana | 0.05 |
| Egg | 2.03 |
| Turkey berries<sup>a</sup> | 0.87 |

<sup>a</sup> not asked about in the FFQ

### Ethics Statement

This study received approval from the UNC Institutional Review Board (IRB) (IRB No. 22-0085). We did not collect biological samples or any other identifying information (in accordance with the HIPAA “Minimum Necessary Rule”) so the risk to participants was minimal. The study received KBTH Scientific and Technical Committee approval, and we obtained formal written consent from the participants.

### Dietary Assessment

Through purposive sampling, clinic participants were approached and consented by 3 RAs, who administered the FFQ in the clinic waiting room with minimal disruptions to the clinic flow prior to each provider visit.

Responses were entered into the REDCap Global survey database as shown in Figure 3 hosted at UNC Chapel Hill.^18,19^ Concentrations of dietary Se in each food item of the FFQ were determined by referencing the standard Se concentration per food unit, determined by the USDA FoodData Central database.^20^ Foods not listed in this database and endemic to Ghana were run as samples through inductively coupled plasma mass spectrometry (ICP-MS) at the Ghana Standard Authority (GSA) as shown in Table 1. Briefly, 1.0g of each sample was weighed and digested using a microwave digester and allowed to cool. The digest was diluted with deionized water. Se concentrations were determined using ICP-MS. Selenium concentration was calculated using the formula below;

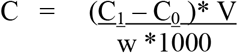

where

C_1_ is the concentration in microgram per litre of the samples

C_0_ is the concentration in microgram per litre of the blank sample

w is the weight of the samples taken

V is the volume of the test sample

### Analysis techniques

As this was a feasibility study without formal hypothesis testing, the analyses were mainly descriptive.^21^ Most of the literature guiding statistical testing in feasibility studies concurs that interpretation and reporting of intervention testing must be done with caution due to the probable small sample size without known power.^21^ Total population Se intake analysis was performed using Stata version 16.0.^22^

## Results

We approached a total of 117 participants attending the clinic over 3 days. We administered 104 total FFQs with an 88.9% recruitment rate. Thirteen women declined to participate for reasons including not speaking English or declining to participate; 1 was in labor (Figure 1). Of the 104 FFQs administered, 100 surveys were eligible to be used in the analysis; 2 were missing, 1 survey was without consent and 1 was missing a page of responses.

**Figure 1.**
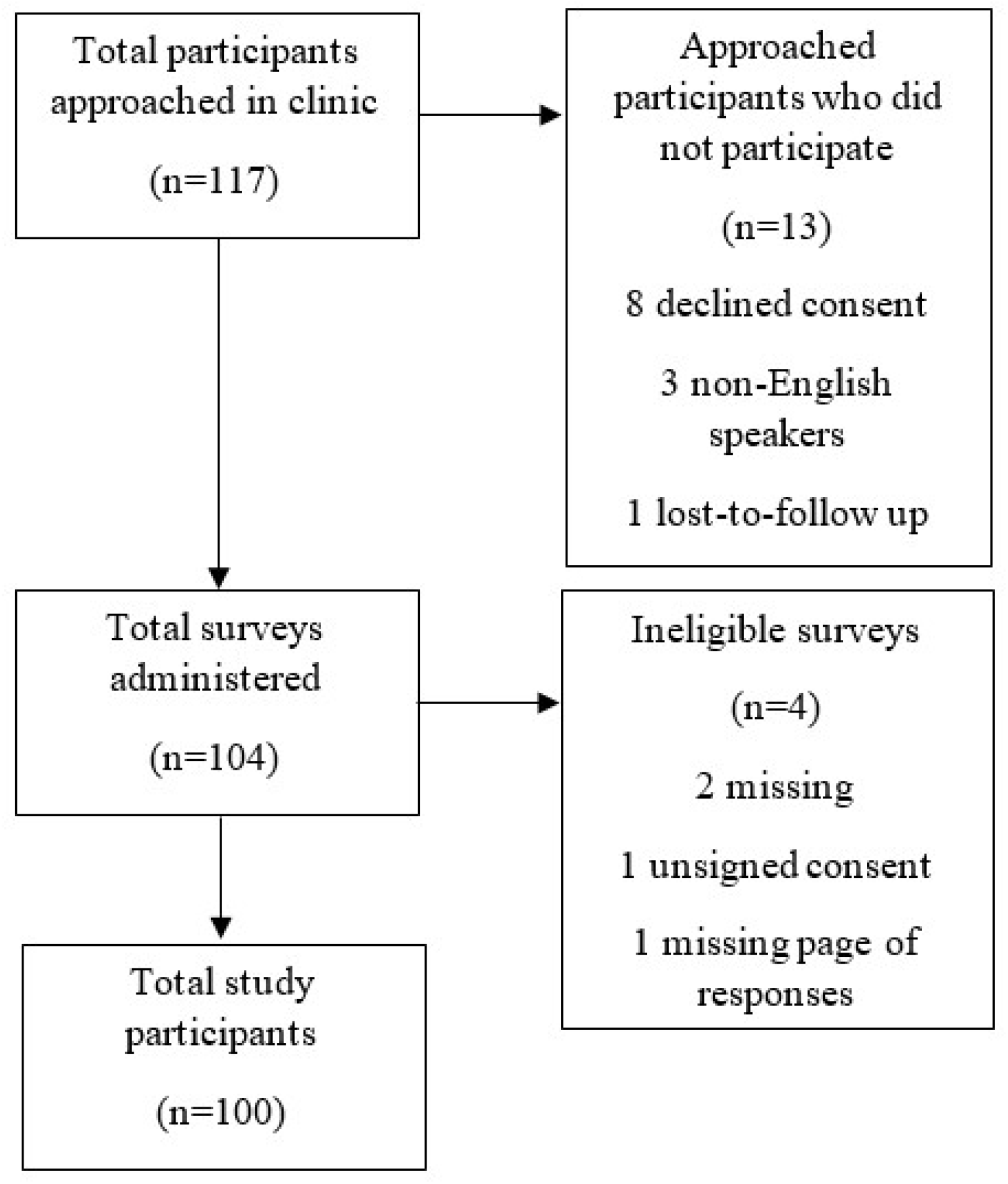
Feasibility study selection CONSORT flow chart.

The median time to complete a single FFQ was 10 minutes, with a range of 7-18 minutes. This time did not include the time taken to administer the consent form, which also varied between RAs. We found that this population of 100 pregnant women had above average Se intake (90.4 μg/day ±50.0 μg).

We found that it was feasible to administer FFQs in the KBTH maternal outpatient waiting room. However, considering the issues of confidentiality when administering the surveys in an open setting, it would be more feasible and appropriate to administer the surveys in a setting with a fixed schedule and within available consulting rooms in which RAs may present the survey to participants while they await their clinician visit.

## Discussion

### On Recruiting English-Speaking Participants

Though English is the national language of Ghana and is taught in primary school education, Akan is most widely spoken with English as the second language. It is important to state the role of the nurse midwives. At the beginning of each day of data collection, the nurse midwives emphasized the importance of speaking “*brofo*”, or “White man’s language”, in Akan, during their daily announcement to the participants. Recruiting English speaking participants in this setting, as a result, was more feasible than we had expected as we reached our maximum study participant goal.

The research team could easily distinguish between participants who had arrived later on in the day and had not heard the midwives’ announcement – this group of pregnant women was more likely to decline participation in the study. We mitigated this by requesting the nurse midwives to repeat the announcement halfway through the day, when most of the initial participants had left.

### On Survey Interpretation and Understanding

The team had post-collection discussions to assess comprehensibility and then identify and address survey issues. For example, we noticed that participants were more familiar with “black-eyed peas” and “Bambara beans” than both “red beans” and “kidney beans”; the latter were listed first.^23,24^ If an RA asked about the less familiar beans first, the general sense was that the respondent was less likely to report that she consumed beans. However, *waakye*, a staple rice and beans dish in the Ghanaian diet, contains black-eyed peas. Presumably, women would be underreporting their intake of beans overall if they did not hear the more commonly consumed “black eyed peas” listed first. Alternatively, this may lead to double counting.^23^ Gerdessen and colleagues optimized the selection of foods for the questionnaire by way of modeling and utilizing a software program to reduce this issue among other errors inherent to FFQ use.^25^ We modified the survey each day to respond to issues like this and refined our survey so that there were no more perceivable corrections to make.

The research team could easily distinguish between patients who had arrived later on in the day and had not heard the midwives’ announcement – this group of pregnant women was more likely to decline participation in the study. We mitigated this by requesting the nurse midwives to repeat the announcement halfway through the day, when most of the initial patients had left.

### Optimal Faculty for FFQ Administration

We found that it was feasible to recruit 6 RAs to administer the FFQ, rotating 2 RAs over the 3 day period, including the PI. As the PI was an “outsider” to the culture, despite the fact that she spoke English, we aimed to primarily use research assistants to carry out the research in order to reduce the gaps in responses and cultural understanding.^26^ Stevano and Deane (2017) highlight some of these gaps as a limited ability to verify the accuracy of the information collected, respondents’ uneasiness to speak to an outsider, etc.

. Our approach of rotating our RAs over each day of data collection introduced the risk of inconsistencies in the nuances of data collection, despite standardized training. We recommend a more reliable and cost-effective method—delegate 2 or 3 RAs at the beginning of the data collection to administer all surveys over the collection period.

### Average FFQ Administration Time

In the context of the waiting room, the average time to administer the 66-itemsurvey was 10 minutes (CI 95%: [9.7, 10.3 minutes]), excluding time to obtain consent. The average time to administer a survey depends on its item number, various studies show. As expected there is a direct correlation between the item number and average time (e.g 14 mins for a 146-item survey).^27^ Initially, we anticipated taking greater than 30 minutes per survey to set up the clinic room before and after each participant encounter and then administer the consent and survey. However the process of administering the FFQ in the waiting room quickened the process. Time taken to obtain consent should be considered, however, as the total time spent with a participant may influence recruitment and engagement.

### On Number of Surveys Per Day

The number of FFQs administered depended on the clinic volume of the day amongst other variables.^24^ We reassessed the initial 5-day data collection period and determined that it was feasible to administer 100 surveys over a period of 3 days. We administered 26, 44, and 34 surveys on the first, second and third days, respectively.

### On Data Management

All 104 paper surveys and informed consent forms were scanned by phone (Adobe Scan) and saved to a secure online folder. Data from all 104 surveys were then manually entered into the REDCap database. It is feasible to use pen and paper in this setting, but we recommend using multiple digital tablets, which could simultaneously record and enter data directly into REDCap or other data collection instruments. Though this study was limited in its financial capacity to procure a handful (2-3) of new study-tablets, the KBTH setting would not prohibit the effective use of such technology as there is secure WiFi and hotspot availability.

### On Data Analysis and Coding

We found that data analysis for our original purposes of determining the average level of Se content in the KBTH population of pregnant women was feasible. The above average Se intake in this population (90.4 μg/day ±50.0 μg) predicts that the rates of documented adverse pregnancy outcomes would be low in this population. Nevertheless, Thompson & Subar (2013), suggest that the error inherent in the FFQ approach renders analysis to approximate quantitative parameters inappropriate; it should be regarded as an estimation if at all. ^23^

### On Appropriate Population-Specific Measures

FFQs primarily include the foods that have high levels of the nutrient of focus and foods that are specific to the context in which they are being administered.^28^ Future studies in the same setting should include foods like oysters and other seafood, which are both known to be high in Se and which pregnant women in Ghana tend to eat in moderation. Consuming snails, however, has long been associated with the taboo of birthing a child who drools excessively.^29^

We recommend creating a dietary survey incorporating the input of women in the community, as this would have not only been helpful but also extraordinarily valuable to the adaptation or validation of the FFQ. During survey administration, researchers will likely gather some information about cultural beliefs and perceptions of food, like the one mentioned above, but formalizing the process through a focus group would serve the research well.

### Strengths and Limitations

The main strength of our study relates to its prospective design which enabled direct data collection and hence minimal recall bias. In addition, this was a feasibility study being the first of its kind in the country. Every limitation presents as a point of edification for a future study design. We made the decision to not collect any protected health information (PHI) in order to maintain confidentiality and adhere to the “Minimum Necessary Rule” of HIPAA.^30^ PHI was not necessary for the purposes of this study that simply assessed feasibility. If PHI had been collected, we would recommend that the results of statistical analysis and intervention testing be interpreted with caution as a power calculation to support the results is not typical of a feasibility study.

Confidentiality remained a limitation of this study as we administered surveys in the waiting room as women arrived for their visits. Though confidentiality may have been compromised in such a setting, we mitigated the risk by not collecting any identifying information in the surveys. Nevertheless, to ensure complete confidentiality in a future study, we recommend collaborating with hospital management and clinicians to use a private clinic room before or after the visit is finished.

## Conclusion

Administration of the Se-focused FFQ to Ghanaian pregnant women attending the antenatal clinic at KBTH is feasible. Data collection ran with minimal to no disruption of the clinic flow. In this setting, where the waiting time was primarily done outside of the clinic room, we had to sacrifice a level of confidentiality that was significantly mitigated by our decision not to collect any identifying information, including demographic information, as enforced by the Minimum Necessary Rule of the IRB. Future studies would do well to incorporate survey validation as well as a context-specific and micronutrient-focused FFQ for effective data analysis. Done effectively, FFQ administration to populations of pregnant women in resource-limited settings, like the one in this study, can improve context-specific prenatal nutritional counseling about Se and, ultimately, improve pregnancy outcomes.

## Data Availability

Data will be made available upon reasonable request.

## Author contributions

AA designed, planned, and conducted the study, completed the data analysis and authored the manuscript. GE and EOA contributed to the design, planning of the study and manuscript writing. MA contributed to the planning and conduct of the study, and then manuscript writing. KAB supervised the design, planning and conduct of the study and contributed to the manuscript writing.

## Acknowledgements

We would like to thank Dr. Tracy Manuck for her support and guidance of this study, which we modeled after her previous work. We would like to thank UNC Gillings School of Public Health for its enablement and support of this project. Lastly, we thank the staff of the KBTH Obstetrics and Gynecology department and the National Service research assistants at NMIMR who were integral in carrying out this study.

## References

1. Darnton-Hill I, Mkparu UC. Micronutrients in pregnancy in low- and middle-income countries. Nutrients 2015; 7: 1744–1768.

2. Stoffaneller R, Morse NL. A review of dietary selenium intake and selenium status in Europe and the Middle East. Nutrients 2015; 7: 1494–1537.

3. Wilson RL, Bianco-Miotto T, Leemaqz SY, et al. Early pregnancy maternal trace mineral status and the association with adverse pregnancy outcome in a cohort of Australian women. J Trace Elem Med Biol 2018; 46: 103–109.

4. Zachariah R, Harries AD, Manzi M, et al. Acceptance of anti-retroviral therapy among patients infected with HIV and tuberculosis in rural Malawi is low and associated with cost of transport. PLoS One; 1. Epub ahead of print 2006. DOI: 10.1371/journal.pone.0000121.

5. Hofstee P, Bartho LA, McKeating DR, et al. Maternal selenium deficiency during pregnancy in mice increases thyroid hormone concentrations, alters placental function and reduces fetal growth. J Physiol 2019; 597: 5597–5617.

6. U.S. Department of Health and Human Services. Selenium Fact Sheet for Health Professionals. NIH Office of Dietary Supplements, https://ods.od.nih.gov/factsheets/Selenium-HealthProfessional/#h2 (2021).

7. Hofstee P, Cuffe JSM, Perkins A V. Analysis of selenoprotein expression in response to dietary selenium deficiency during pregnancy indicates tissue specific differential expression in mothers and sex specific changes in the fetus and offspring. Int J Mol Sci 2020; 21: 1–19.

8. Lewandowska M, Sajdak S, Lubiński J. The role of early pregnancy maternal selenium levels on the risk for small-for-gestational age newborns. Nutrients; 11. Epub ahead of print 2019. DOI: 10.3390/nu11102298.

9. Perkins A V., Vanderlelie JJ. Multiple micronutrient supplementation and birth outcomes: The potential importance of selenium. Placenta 2016; 48: 61–65.

10. Guo X, Zhou L, Xu J, et al. Prenatal Maternal Low Selenium, High Thyrotropin, and Low Birth Weights. Biol Trace Elem Res 2021; 199: 18–25.

11. Lewandowska M, Lubiński J. Serum microelements in early pregnancy and their risk of large-for-gestational age birth weight. Nutrients 2020; 12: 1–16.

12. Sjöström L, Peltonen M, Jacobson P, et al. Bariatric surgery and long-term cardiovascular events. JAMA - J Am Med Assoc 2012; 307: 56–65.

13. Laar A., Ampofo W., Tuakli J, et al. Factors associated with suboptimal intake of some important nutrients among HIV-positive pregnant adolescents from two Ghanaian districts. J Ghana Sci Assoc; 11. Epub ahead of print 2010. DOI: 10.4314/jgsa.v11i2.50930.

14. Eze SC, Ododo NA, Ugwu EO, et al. Serum selenium levels of pre-eclamptic and normal pregnant women in Nigeria: A comparative study. PLoS One 2020; 15: 1–13.

15. Asare GA, Nani A. Serum levels of Cu, Se, and Zn in adult rural/urban residents in Ghana: Paradigm shift? Biol Trace Elem Res 2010; 137: 139–149.

16. Holmquist E, Brantsæter AL, Meltzer HM, et al. Maternal selenium intake and selenium status during pregnancy in relation to preeclampsia and pregnancy-induced hypertension in a large Norwegian Pregnancy Cohort Study. Sci Total Environ 2021; 798: 149271.

17. http://KBTH.gov. Obstetrics & Gynaecology Department – Korle-Bu Teaching Hospital, https://kbth.gov.gh/departments-centres/obstetrics-gynaecology-department-2/ (accessed 2 April 2022).

18. Harris P, Taylor R, Minor B, et al. EDCap Consortium, The REDCap consortium: Building an international community of software partners. J Biomed Inform. Epub ahead of print 2019. DOI: doi: 10.1016/j.jbi.2019.103208].

19. Harris P, Taylor R, Thielke R, et al. Research electronic data capture (REDCap) – A metadata-driven methodology and workflow process for providing translational research informatics support,. J Biomed Inf 2009; 42: 377–81.

20. http://Usda.gov. U.S Department of Agriculture Food Data Central. n.d., https://fdc.nal.usda.gov/.

21. Arain M, Campbell MJ, Cooper CL, et al. What is a pilot or feasibility study? A review of current practice and editorial policy. BMC Med Res Methodol 2010; 10: 1–7.

22. StataCorp. Stata Statistical Software: Release 16.

23. Thompson FE, Subar AF. Dietary assessment methodology. 2013. Epub ahead of print 2013. DOI: 10.1016/B978-0-12-802928-2.00001-1.

24. Molag ML, De Vries JHM, Ocké MC, et al. Design characteristics of food frequency questionnaires in relation to their validity. Am J Epidemiol 2007; 166: 1468–1478.

25. Gerdessen JC, Souverein OW, Van ‘T Veer P, et al. Optimising the selection of food items for FFQs using Mixed Integer Linear Programming. Public Health Nutr 2015; 18: 68–74.

26. Stevano S, Deane K. The role of research assistants in qualitative and cross-cultural social science research. Handb Res Methods Heal Soc Sci 2019; 1675–1690.

27. Franco RZ, Alawadhi B, Fallaize R, et al. A Web-based graphical food frequency assessment system: Design, development and usability metrics. JMIR Hum Factors; 4. Epub ahead of print 2017. DOI: 10.2196/humanfactors.7287.

28. Shim J-S, Oh K, Kim HC. Dietary assessment methods in epidemiologic studies. Epidemiol Health 2014; e2014009.

29. Senah K. MATERNAL MORTALITY IN GHANA : THE OTHER SIDE 1 Kodjo Senah Abstract. Mortality 2003; 1: 47–55.

30. HHS Office for Civil Rights. OCR HIPAA Privacy MINIMUM NECESSARY [45 CFR 164.502(b), 164.514(d)]. Heal Inf Priv 2003; 502: 2002–2004.

